# CDS-PD: A Novel Clinical Decision Support Platform for Parkinson’s Disease

**DOI:** 10.1101/2025.07.24.25331875

**Authors:** Deepak K. Gupta, Pedram Golnari, Katrina Prantzalos, Ian Zurlo, Vivikta Iyer, Brenna M. Lobb, Cole Zweber, Manu Bulusu, Dakota Clarke, James T. Boyd, Curtis Tatsuoka, Amie L. Hiller, Satya S. Sahoo

## Abstract

The Movement Disorders Society clinical diagnostic criteria for Parkinson’s disease (MDS-PD) allow highly sensitive and specific diagnosis of Parkinson’s disease. However, their adoption has been limited due to lack of a clinical decision support (CDS) tool to support clinicians and researchers in systematically and accurately applying the MDS-PD criteria. We have developed and performed preliminary validation of a CDS platform for PD (CDS-PD) as a modular and extensible informatics platform with comprehensive functionalities for recording relevant patient information. We have performed real-time application of diagnostic algorithm of the MDS-PD criteria. The CDS-PD platform shows high concordance with application of the MDS-PD criteria by experienced movement disorders neurologists for established PD (disease duration ≥5 years). The CDS-PD platform is a step towards realizing the standardized electronic implementation of the MDS-PD criteria for PD patient care and clinical trials at point-of-care. The CDS-PD platform can be accessed after registration at weblink https://www.cdspd.org.

## INTRODUCTION

Parkinson’s disease (PD) is the second most common neurodegenerative disorder after Alzheimer’s disease[1] and it is the fastest growing neurological disorder globally in terms of prevalence and disability[2]. There is an increasing concern in the healthcare community that these worldwide prevalence trends point to a potential PD epidemic[3]. Therefore, an early and accurate diagnosis of PD is crucial for both clinical care and research using systematic application of validated formal diagnostic criteria. Specifically, the accuracy of clinical diagnosis without the application of diagnostic criteria can be low in primary care and even in general neurology clinics where movement disorders specialty expertise may not be readily available[4]. Achieving a high accuracy of diagnosis is particularly important as neuroprotective clinical trials[5] and preventative strategies[6] focus on early-stage PD[5–8], which is especially challenging to differentiate from related atypical parkinsonian disorders (APD), namely dementia with Lewy bodies[9], multiple system atrophy[10], progressive supranuclear palsy[11], and corticobasal degeneration[12].

The development of the International Parkinson and Movement Disorder Society (MDS) Clinical Diagnostic Criteria for Parkinson’s disease (MDS-PD Criteria)[13] represents a critical step forward towards aiding accurate early diagnosis of PD in clinical care and trials. Since their publication over a decade ago in 2010, the MDS-PD criteria have been validated by expert neurologists with 10 years or more of experience in PD[14] and against post-mortem neuropathological diagnosis[15], and thus considered global standard for PD diagnosis both for clinical and research use[16]. However, applying the MDS-PD criteria using the existing paper-based format requires the neurologists to address two key challenges: (1) capturing a diverse set of patient information, including time dependent variables, with consistent quality; and (2) manually applying a complex, multi-step algorithm to generate an accurate diagnosis. In context of universal adoption of the electronic health record (EHR) systems in the United States[17], the current lack of support for the MDS-PD criteria in leading EHR systems has severely limited their adoption in real-world clinical practice and research. Thus, there is an unmet need to develop clinical decision support (CDS) to support the neurologist in applying the MDS-PD criteria at point-of-care.

We address this above gap through the first CDS platform for PD (CDS-PD) that implements the MDS-PD criteria using an advanced informatics software stack and a distributed deployment architecture for supporting multicenter research study. The CDS-PD platform comprehensively addresses the two integral components of CDS: (1) accurate capture of patient’s information through a neurologist designed and structured user interface intended to represent existing clinical workflows; and (2) real-time application of algorithm on the recorded information to perform diagnostic classification on the MDS-PD criteria. Here, we report the development and preliminary validation of the CDS-PD platform for established PD (disease duration ≥5 years).

## OBJECTIVES

The CDS-PD platform realizes the potential of the MDS-PD criteria in enhancing early and accurate diagnosis of PD patients at the point of care. It is built for multi-institution, distributed deployment with modular design to evolve with changing diagnosis guidelines and clinical needs of PD patients.

## METHODS

### MDS-PD Criteria

The MDS-PD criteria[13] consist of four categories of different criteria and an algorithm: essential criteria (*which is a prerequisite*), absolute exclusion criteria and red flags criteria (*which collectively constitute negative features*) and supportive criteria (*which constitute positive features*); the algorithm involves establishing essential criteria, ruling out absolute exclusion criteria, and with weighing/counterbalancing of red flags criteria with supportive criteria to allow classification of the patient into two distinct levels of diagnostic certainty for PD, namely, “*Clinically Established PD*”, “*Clinically Probable PD”*, or “*Not PD*” (figure 1). Of note, the MDS-PD criteria can only be operationalized on the patient information (history, neurological examination, and relevant diagnostic tests) captured by a clinician with sufficient training in neurology[16].

**Figure 1:**
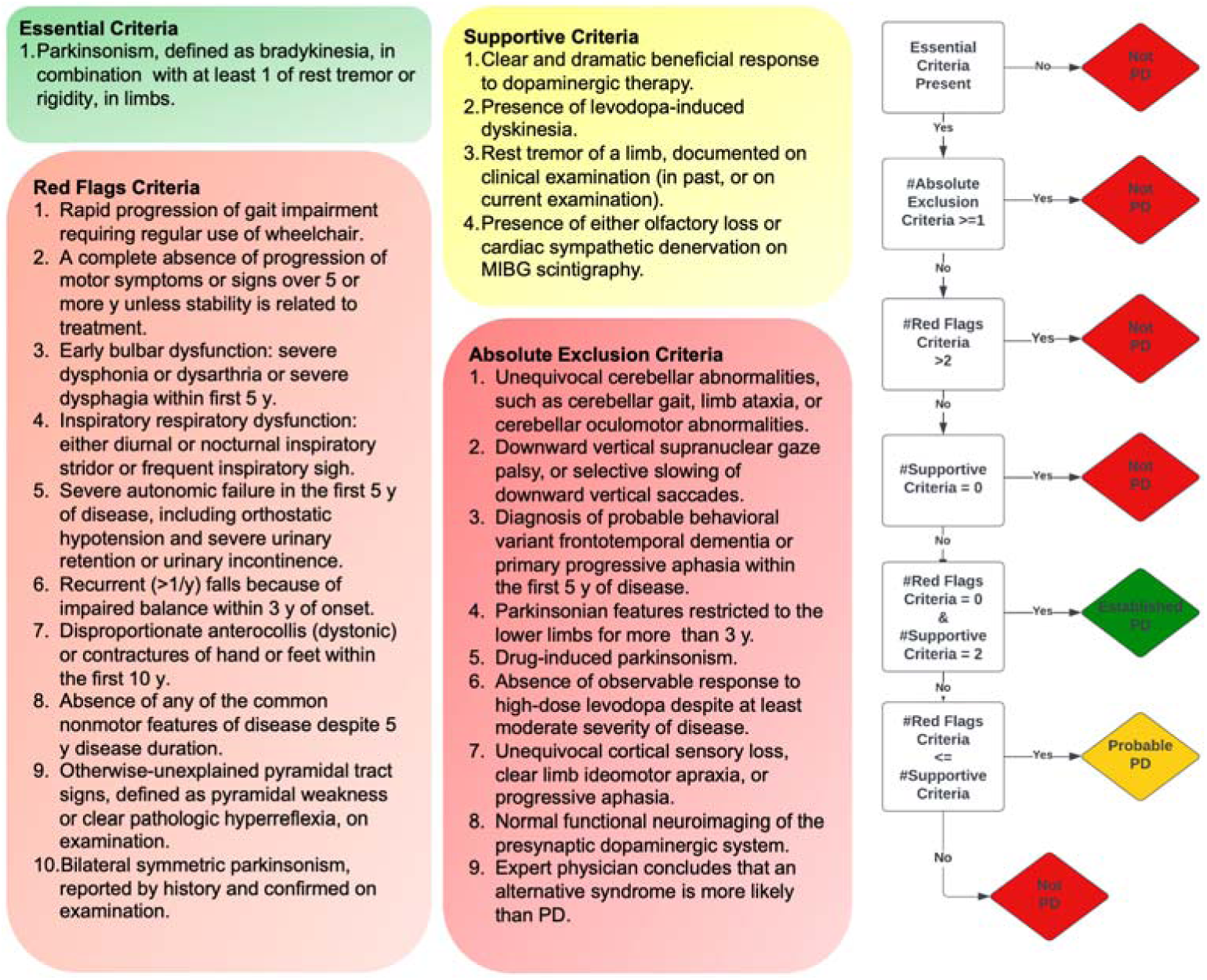
Four categories of criteria and algorithm of the MDS-PD criteria.

### CDS-PD Software Development

The CDS-PD platform incorporates an extensible design framework with integrated modules for data recording and analysis features that are implemented using the Django web development software stack[18]. The Django software uses a Model-View-Controller (MVC) architecture with the Model storing the data in an object-relational model, the View encoding the user interface, and the Controller encoding the functionalities of the application, including user interactions. The CDS-PD platform uses PostgreSQL relational database management system to manage data and incorporates a role-based access control (RBAC) module to support user accounts and access. The CDS-PD platform is accessible as a browser-based web application with support for multi-institution clinical research studies.

This platform was developed following an agile, iterative software engineering methodology. Each development cycle consisted of requirement gathering from movement disorder neurologists, prototype implementation, internal quality assurance testing, and structured user acceptance testing (UAT) sessions. Version control was maintained using Git with a protected main branch, feature-specific development branches, and formal release notes. A multi-layer quality assurance process included automated unit tests for algorithmic logic, integration testing for database queries, and regression testing after each deployment.

### Software Quality Assurance & User Acceptance Testing

We systematically evaluated the CDS-PD platform as part of a prospective clinical research study investigating diagnostic and prognostic differences between PD patients with or without history of exposure to agent orange. This multicenter study is approved by the single institutional review board (IRB), WCG IRB, and is being conducted at the University of Vermont Medical Center (UVMMC), the Oregon Health & Sciences University (OHSU), and the Veteran Affairs Portland Healthcare System (VAPORHCS), with grant funding from the Congressionally Directed Medical Research Programs (CDMRP) through the Department of Defense (DoD)[19].

Software quality assurance (QA) procedures included automated validation of criterion-specific logic, stress testing of PostgreSQL query performance, and manual semantic verification of algorithmic outputs. UAT was conducted with neurologists and research coordinators at UVMMC, OHSU, and VAPORHCS. These UAT sessions identified workflow bottlenecks, ambiguous field descriptions, and UI improvements, all of which were incorporated into iterative platform updates.

### Data Collection Protocol

Patients were recruited from each site after they had signed a written IRB-approved consent form. Neurologists entered examination components requiring clinical expertise, including the MDS-UPDRS Part III and neurological examination findings. Research coordinators entered demographic, historical, diagnostic, and medication data. Missing data were minimized by mandatory-field validation and database-level constraints. No required MDS-PD fields were left incomplete. The study did not perform formal inter-rater reliability testing because each patient was evaluated by only one neurologist per visit.

### Validation Study Protocol

Clinician assessments were completed first on paper during the research encounter. Platform processing occurred only after full data entry, ensuring clinicians were blinded to CDS-PD results. The platform’s classifications were compared to clinician diagnoses. Discordant cases were adjudicated by reviewing structured symptom entries, which revealed that inspiratory stridor (Red Flag #4) was documented but unintentionally overlooked during paper-based review. There were no cases where the platform was unable to make an assessment due to incomplete criteria, as all 43 patients had complete data.

The validation study followed a structured protocol aligned with STARD-AI[20] and STARE-HI[21] recommendations. The timing of assessments, blinding procedures, adjudication workflow, and data completeness requirements were prospectively defined. Platform feasibility, diagnostic performance, and edge-case behavior (e.g., ambiguous findings or conflicting exam inputs) were systematically recorded. No cases failed classification due to incomplete required data.

### Algorithm Implementation

All 24 criteria from the MDS-PD framework were translated into Boolean logic mapped to database variables. Ambiguous clinical findings invoked confirmation prompts to ensure fidelity. Missing or inconsistent data prevented algorithm execution until resolved. Implementation fidelity was validated using synthetic test cases and expert clinician review. Full pseudocode of all criteria are provided in the Supplementary Appendix.

## RESULTS

The CDS-PD platform currently consists of two modules: (1) Patient Information Recording Module; and (2) Patient Information Analysis Module. The former module allows systematic and detailed entry of patient information while ensuring data quality rules, while the latter allows subsequent automated application of algorithm of the MDS-PD criteria (as detailed below and depicted in a video-walkthrough of this module provided in supplementary material 1).

### Patient Information Entry Module

This module closely models clinical workflow typically used by movement disorders specialists for evaluation of a patient for PD diagnosis using the MDS-PD criteria. It is implemented using a tab-structured interface consisting of 11 sections/tabs: *demographics, motor symptoms, non-motor symptoms, family history, social & environmental history, occupational & military history, health profile, medications, exam, scales, domain-specific information (DSI), and diagnostics.* These sections also incorporate the National Institute of Health (NIH) Common Data Elements (CDE), for example, family history CDE for Parkinson’s disease[22].

Each information category/tab is further organized into appropriate subsections/subtabs corresponding to the required level of granularity. For example, the Exam section/tab is organized into five subsections: *General Exam, Neurological Exam, Basic Movement Disorders Exam, Additional Movement Disorders Exam, and Advanced Movement Disorders Exam* (figure 2). The Basic Movement Disorders exam subsection/subtab captures the patient information corresponding to the most fundamental motor examination features of PD, specifically, parkinsonism (bradykinesia, rigidity, rest tremor), action tremor (postural, kinetic tremor), and dyskinesia.

**Figure 2:**
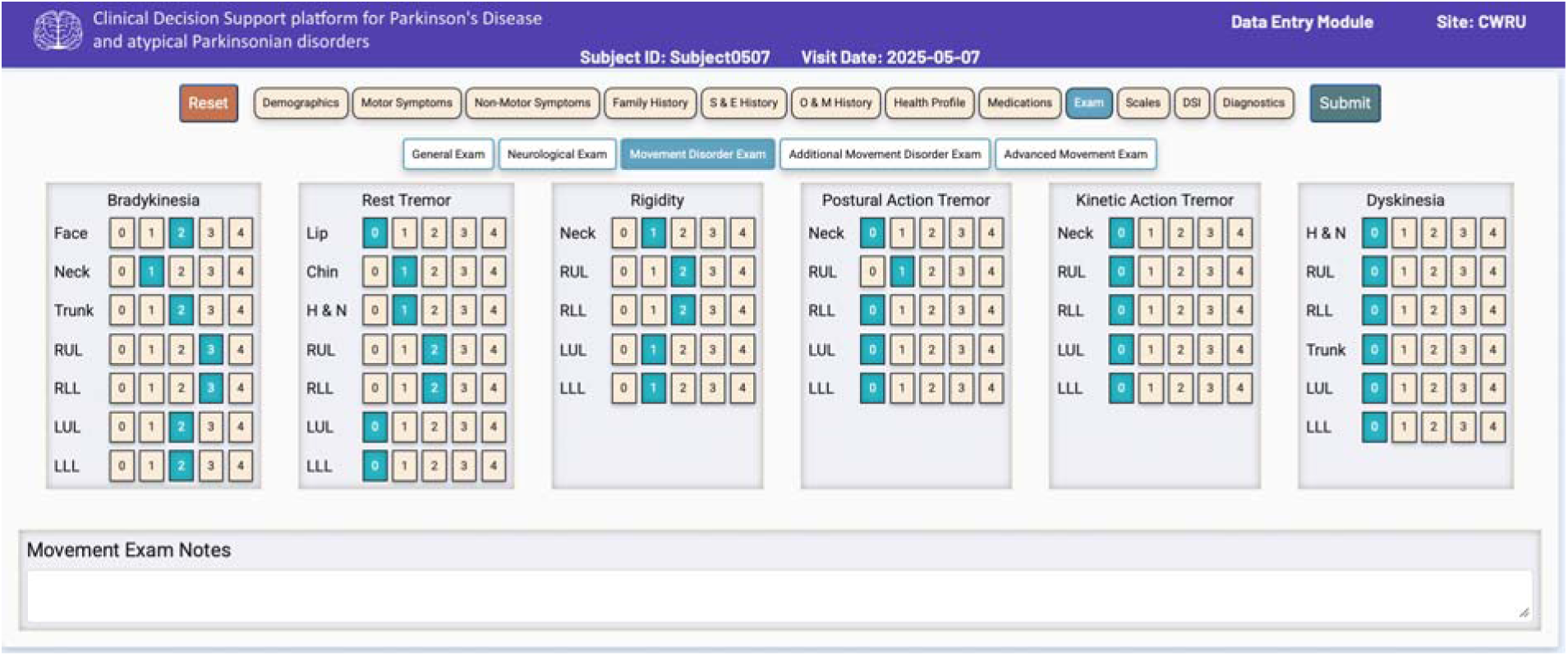
A screenshot of the Patient Information Entry Module depicting 11 data sections/tabs and “Movement Disorders Exam” subsection of the “Exam” data section/tab. *Note: Subject0507 shown in figure is a dummy/test subject*.

The user interface (UI) consists of intuitive selection buttons, drop-down menus, calendar dates, and in a few cases free text fields. The UI incorporates jQuery validation libraries and rule-based validation checks for: (1) correct data type for a data entry field, (2) range value: to verify the validity of values within predefined limits for a given data element, and (3) value presence: to ensure that mandatory fields are completed by the user. These built-in data quality checks reduce the occurrence of missing information as well as incorrect data entries with minimal additional burden on users.

### Patient Information Analysis Module

This module implements the four categories of criteria (essential, absolute exclusion, supportive, red flags) and the algorithm of the MDS-PD criteria as series of queries to the PostgreSQL database management system (DBMS) and leverages the DBMS query optimizer for fast execution of the classification algorithm. Specifically, each of the criteria under the four categories are subdivided into components and subcomponents (as applicable) and mapped to corresponding patient information variables stored in the database, followed by analysis of the variable values according to the condition/logic of the criteria. For example, parkinsonism is the only essential criteria category that consists of three components (*bradykinesia, rigidity, and rest tremor*). These components are first assessed in terms of body part (*upper limb, lower limb, neck*) and laterality (*right or left; for limb only*) as part of the Movement Disorder Society-Unified Parkinson’s Disease Rating Scale (MDS-UPDRS)[23] and then evaluated to be true or false on the condition/logic of “*bradykinesia must be present with or without rigidity or rest tremor*” (figure 3). The rest of the 23 criteria under the other three categories have been similarly implemented.

**Figure 3:**
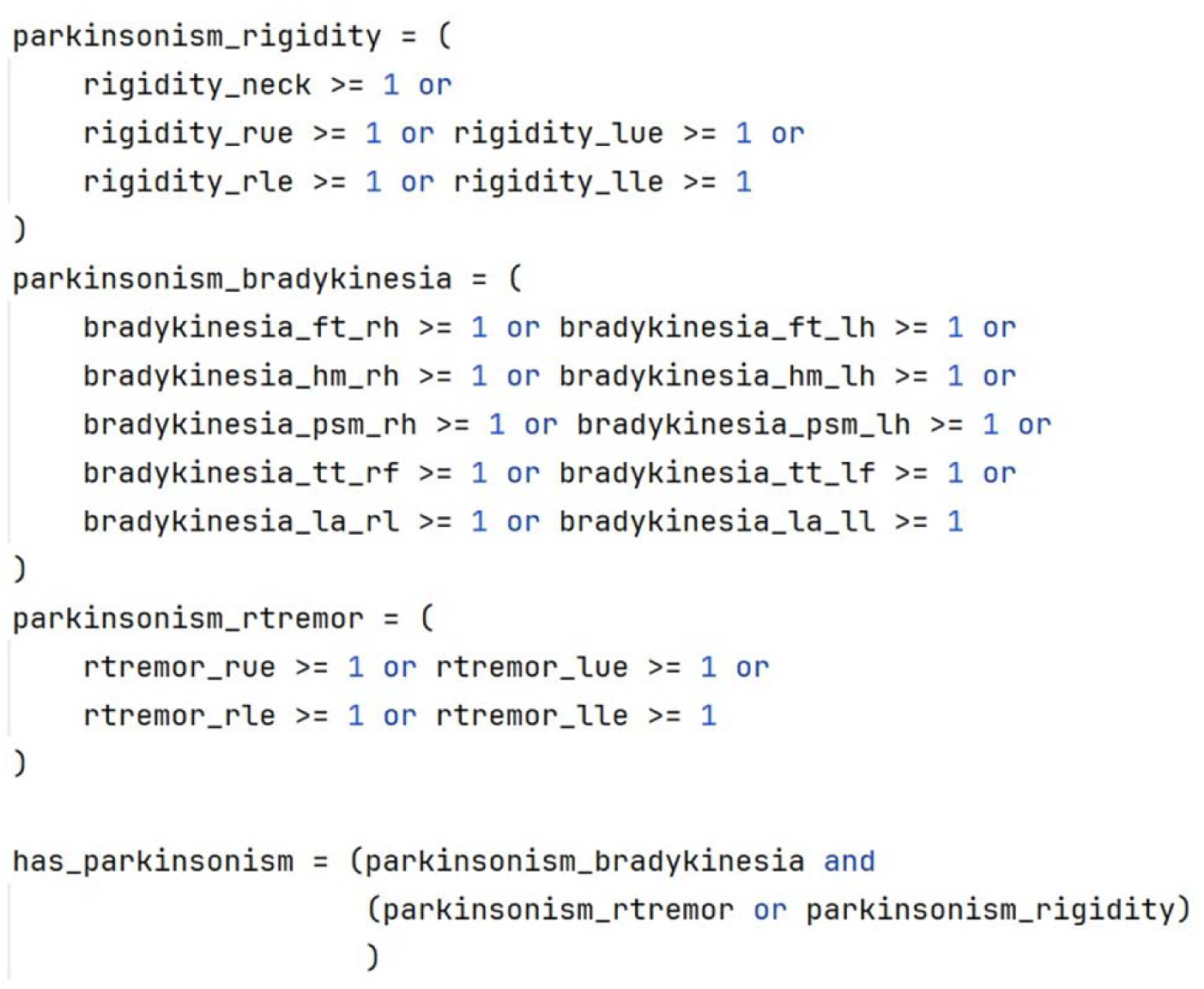
Pseudocode showing implementation of the essential criteria of the MDS-PD criteria.

Once all 24 criteria have been evaluated to a Boolean value (true or false) using the recorded patient information, corresponding values are then evaluated on the algorithm of the MDS-PD criteria to classify the two distinct levels of diagnostic certainty for PD, namely, “*Clinically Established PD*”, “*Clinically Probable PD”*, or “*Not PD*”. For example, the “Clinically Established PD” level of diagnostic certainty requires the following: presence of requisite essential criteria, absence of negative features (all 9 absolute exclusion criteria and 10 red flags), and presence of 2 positive features (supportive criteria) (figure 4).

**Figure 4:**
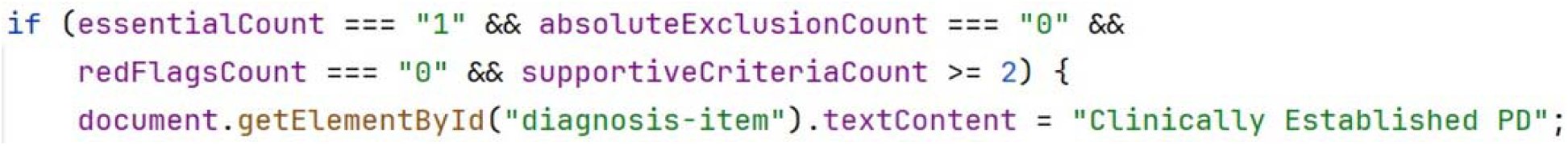
Algorithm for “Clinically Established PD” diagnostic level of the MDS-PD criteria.

### Evaluation

The deployment of the CDS-PD platform in the clinical research study involved the creation of standalone instances of the platform that were deployed on a touchscreen laptop (Microsoft Surface) at the UVMMC site and OHSU/VAPORHCS site. The study personnel (movement disorders neurologists and research coordinators) at both sites were trained in the use of CDS-PD platform with a user guide document and as part of regular virtual team meetings for eliciting their feedback during its development.

The CDS-PD platform was evaluated using a subset (n = 43) of study cohort (PD patients with disease duration >=5 years; table 1) against the standard of clinical diagnosis made by fellowship trained movement disorders neurologists (with over 4 years of independent practice experience at a minimum) using paper-based format of the MDS-PD criteria. Based on this evaluation, the concordance rate of the CDS-PD platform was 91.30% (21 out of 23 matched cases) for the UVMMC site, and 95.00% (19 out of 20 cases) for the OHSU/VAPORHCS site, for an overall concordance rate of 93.02% (95% CI, 81.4% to 97.6%) for the two sites combined using a Wilson Score interval.

**Table 1:** Baseline demographics and clinical characteristics of the Parkinson’s disease patients at the two sites. (MOCA – Montreal Cognitive Assessment, UPSIT – University of Pennsylvania Smell Identification Test).

| Variable | UVMC Site (n=23) | OHSU/VAPORHCS Site (n=20) | All Patients (n=43) |
| --- | --- | --- | --- |
| Age (mean, years) | 70.57 | 75.90 | 73.05 |
| Sex (female/male) | 7 / 16 | 4 / 16 | 11 / 32 |
| Ethnicity (Not Hispanic or Latino) | 22 | 19 | 41 |
| MDS-UPDRS (mean) |  |  |  |
| - Part I | 10.52 | 14.75 | 12.49 |
| - Part II | 16.00 | 20.25 | 17.98 |
| - Part III | 38.09 | 43.65 | 40.67 |
| - Part IV | 1.17 | 5.70 | 3.28 |
| MOCA (mean) | 25.91 | 23.90 | 24.98 |
| UPSIT (mean) | 19.13 | 16.20 | 17.77 |

There were 3 cases of mismatch, 2 at the UVMMC site and 1 at VAPORCHS site. In all 3 of the mismatched cases, the CDS-PD platform classified the patients as “Clinically Probable PD” due to the presence of one red flag (*#4: Inspiratory respiratory dysfunction: either diurnal or nocturnal inspiratory stridor or frequent inspiratory sigh*), while the movement disorders neurologist classified the patients as “Clinically Established PD”. These mismatched cases were identified when comparing the clinician diagnoses with the platform classifications. Adjudication demonstrated that inspiratory stridor had been captured during structured symptom review but was not incorporated into the final paper-based classification. When these data were algorithmically applied, the CDS-PD output reflected the criteria as specified. One potential reason for this recall bias on the part of the movement disorders neurologists might be that inspiratory stridor is a rare motor symptom encountered in PD and anticipated to be present infrequently even in APD.

## DISCUSSION

Despite publication of the MDS-PD criteria a decade ago, their usage has been limited at point of care in clinical practice despite becoming standard of care in PD clinical trials even in their current paper-based format. The lack of CDS approach for the MDS-PD criteria in context of EHR systems is likely a major factor contributing to this gap. Additionally, standardized and electronic application of the MDS-PD criteria through the CDS-PD platform can provide consistent and efficient application of objective diagnostic criteria and thus contribute to increasing the accuracy of PD diagnosis in both clinical care and research compared to non-criteria-based diagnosis. For example, absolute exclusion criteria #1 (*Unequivocal cerebellar abnormalities*), involves various clinical elements of cerebellar gait, limb ataxia, and specific oculomotor abnormalities—e.g., sustained gaze-evoked nystagmus, macro square wave jerks, and hypermetric saccades. Notably, hypermetric saccades, can be observed in advanced PD[24], and thus complicate the differentiation of cerebellar involvement from basal ganglia dysfunction in terms of consistent application of absolute exclusion criteria in a clinically relevant context. These nuances underscore the need for a CDS approach in applying such clinically complex diagnostic criteria. In contrast to manual paper-based application of the MDS-PD criteria, the CDS-PD platform has several inherent advantages: systematic and less error-prone collection of the input data needed on all four categories of criteria included in MDS-PD criteria; computerized modelling of the diagnostic algorithm of the MDS-PD criteria; and computer-assisted processing of the collected input data on the diagnostic algorithm for classifying the diagnostic level of the patient’s diagnosis.

The present study reports preliminary validation of the CDS-PD platform using a cohort of patients with established PD. While this represents only a specific subset of the greater spectrum of parkinsonian disease, the strong degree of concordance using this quintessential cohort lends credence towards potential expansion of the platform to other populations, such as early-stage PD, and the APD. We have since developed the CDS-PD platform to automatically apply the Movement Disorder Society Criteria for Clinically Established Early Parkinson’s Disease[25] for patients with disease duration < 5 years (supplementary material 2).

Furthermore, the CDS-PD platform now also allows for automatic characterization of the PD into clinically relevant subtypes, for example, tremor-dominant (TD) and postural instability and gait dysfunction (PIGD), and indeterminate (IT) subtypes based on patient data derived from the MDS-UPDRS[23] (supplementary material 3). A module tailored to delineating APD is likewise in development for the CDS-PD platform. These differential diagnostic functions will require independent validation using a heterogenous cohort containing both early and established PD, APD, and PD-mimics.

There are notable limitations to this study. From an experimental design perspective, the study is underpowered: with a convenience sample of 43 patients from two sites, validation only for established PD patients, and no negative controls, this limits the generalizability of our findings. This lack of generalizability extends to current platform use in differential diagnostic settings, such in the case of the APD and PD-mimics. These are all limitations that we hope to address with continued development of both additional platform modules, and expansion of our study to more sites and patient cohorts. Additionally, the current study compares the platform’s application of the MDS-PD criteria with paper-based application of the same criteria, and while this is necessary to establish a baseline for simple manipulation of clinical patient data, it does not directly compare to an independent gold-standard of diagnosis such as post-mortem biopsy. However, the MDS-PD criteria have been extensively validated against neuropathological diagnosis. From a developmental design perspective, the CDS-PD platform currently lacks reportable usability metrics. While this is less relevant to the execution of the current study, future expansion and adoption of the platform will necessitate greater internal measurement of the quality and clarity of the user experience. Similarly, the platform currently requires structured data entry by trained personnel (either the neurologist or the site investigator). The platform is being developed with the eventual capability to populate from within the electronic health record, and so this will be addressed in future deployments. Lastly, we acknowledge the limitation that we do not currently have assessment of long-term cost-effectiveness, reliability, or consistency across users, for which evaluation of all will require longitudinal adoption of the platform.

A key aspect of the CDS-PD platform is its modular and scalable architecture that enables it to be expanded to support additional use cases, for example, currently supporting a multicenter project developing novel paradigm of studying cognitive dysfunction against conventional schemes of mild cognitive impairment and dementia in PD[26]. In the short term, we plan to develop the CDS-PD platform for computer-assisted differential diagnosis of PD from APD for general neurologists, using practical algorithms[7, 8, 27], and for movement disorders neurologists, using specific criteria[9–12]. In the long term, we envision that the CDS-PD platform will be interoperable with EHR using SMART (Substitutable Medical Applications, Reusable Technologies) on the FHIR (Fast Healthcare Interoperability Resources) technology framework[28], as well as advance precision medicine for individual PD and APD patients[29, 30] through AI-based data mining of relevant data repositories[31, 32] at point-of-care.

## Data Availability

The CDS-PD platform can be accessed after registration at weblink https://www.cdspd.org

## AUTHOR CONTRIBUTIONS

Deepak Gupta: Conceptualization, Validation, Visualization, Investigation, Data Curation, Software, Formal Analysis, Project Administration, Writing – Original Draft, Writing – Review and Editing

Pedram Golnari: Data Curation, Methodology, Formal Analysis, Software, Writing—Review and Editing

Katrina Prantzalos: Methodology, Software

Ian Zurlo: Conceptualization, Methodology

Vivikta Iyer: Investigation, Data Curation, Formal Analysis, Writing – Review and Editing Brenna M Lobb: Investigation, Data Curation, Formal Analysis, Writing – Review and Editing Cole Zweber: Formal Analysis, Visualization, Writing—Review and Editing

Manu Bulusu: Software

Dakota Clarke: Investigation, Data Curation

James T. Boyd: Writing—Review and Editing

Curtis Tatsuoka: Formal Analysis

Amie L. Hiller: Conceptualization, Methodology, Writing—Review and Editing

Satya S. Sahoo: Conceptualization, Validation, Visualization, Investigation, Data Curation, Formal Analysis, Project Administration, Writing – Original Draft, Writing – Review and Editing

## DATA & CODE AVAILABILITY

The dataset used in this publication will be made available by request from any qualified investigator. To make such request, please contact corresponding author of this paper from your institutional email address.

Source code is not publicly available due to institutional restrictions, but access may be granted to qualified researchers for non-commercial academic use upon reasonable request (subject to data security review and data use agreement). The live platform is accessible to qualified researchers for non-commercial academic use at cdspd.org.

