## Supplementary Appendix for "CDS-PD: A Novel Clinical Decision Support Platform for Parkinson’s Disease"

Supplementary Material 1:

[Video file at OneDrive link](https://uvmoffice-my.sharepoint.com/personal/dkgupta_uvm_edu/_layouts/15/stream.aspx?id=%2Fpersonal%2Fdkgupta%5Fuvm%5Fedu%2FDocuments%2FResearch%20Projects%2FORMIS%2DPD%20DOD%20EIRA%20Project%2FCommon%20Folder%2FPapers%2C%20Presentation%20%26%20Abstracts%2F2023%2D09%20ORMIS%2DPD%20Diagnosis%20Paper%2FJAMIA%20SUBMISSION%20FOLDER%2FSUPPLEMENTARY%20MATERIALS%2FSupplementary%20Material%201%2Emp4&ga=1&referrer=StreamWebApp%2EWeb&referrerScenario=AddressBarCopied%2Eview%2E24dd845b%2D4361%2D474d%2Da49a%2Db10a3bd86e39)

Supplementary Material 2

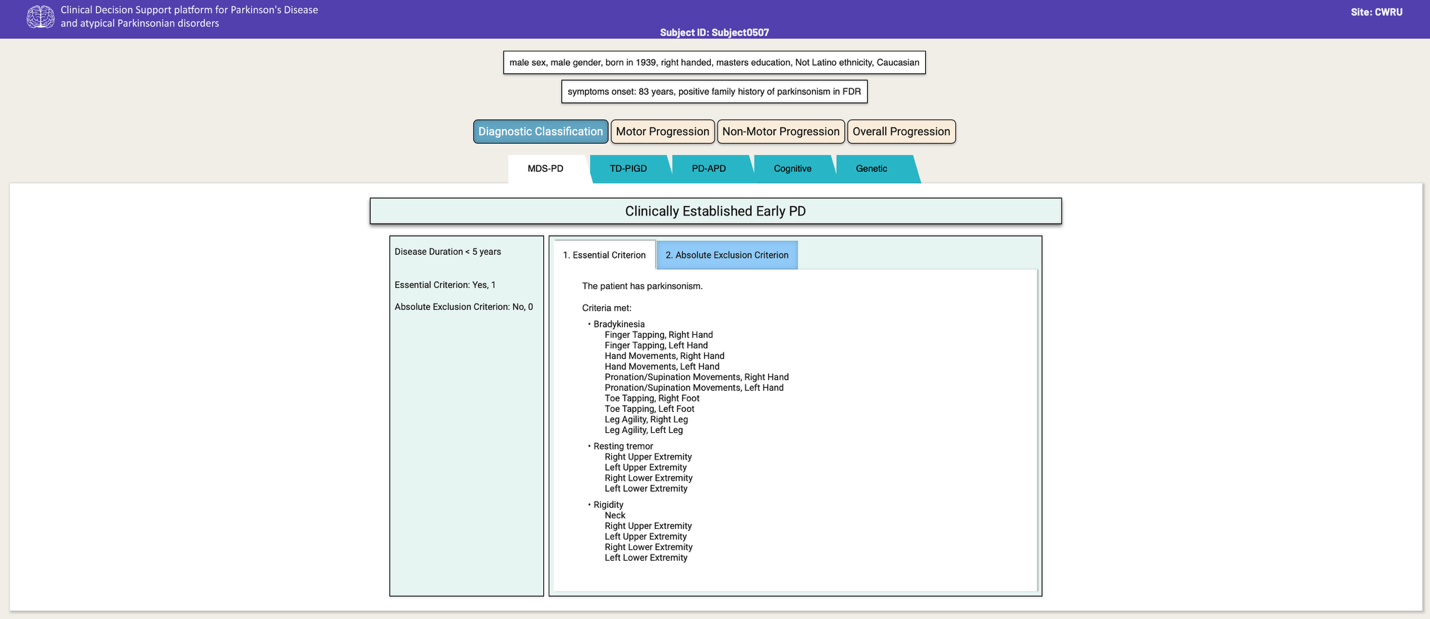

Supplementary Material 3

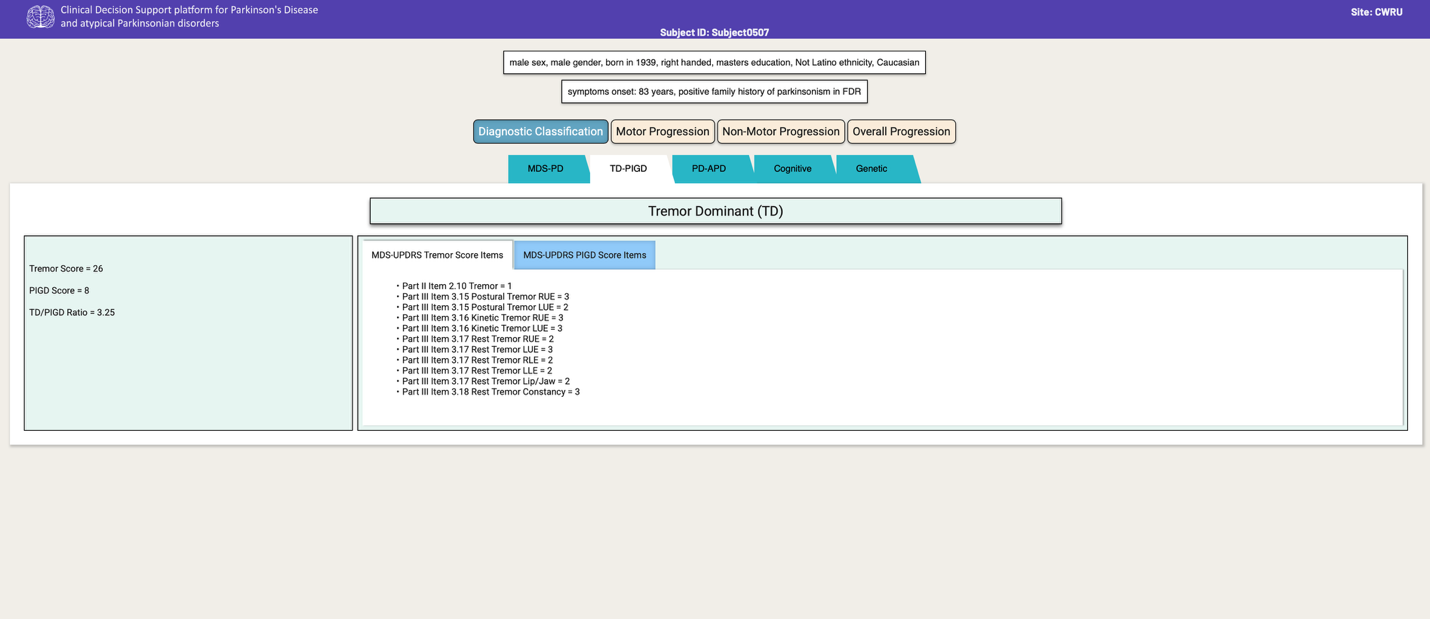

*Note: Subject Test0507 shown in figure 2 and supplementary material is a dummy/test subject.*

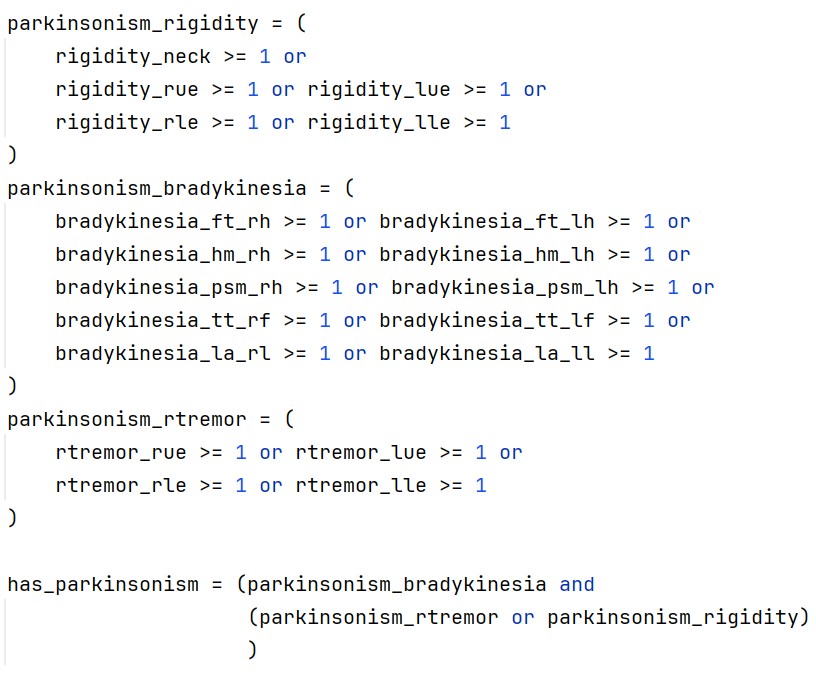
Supplementary Material 4a – Essential criteria

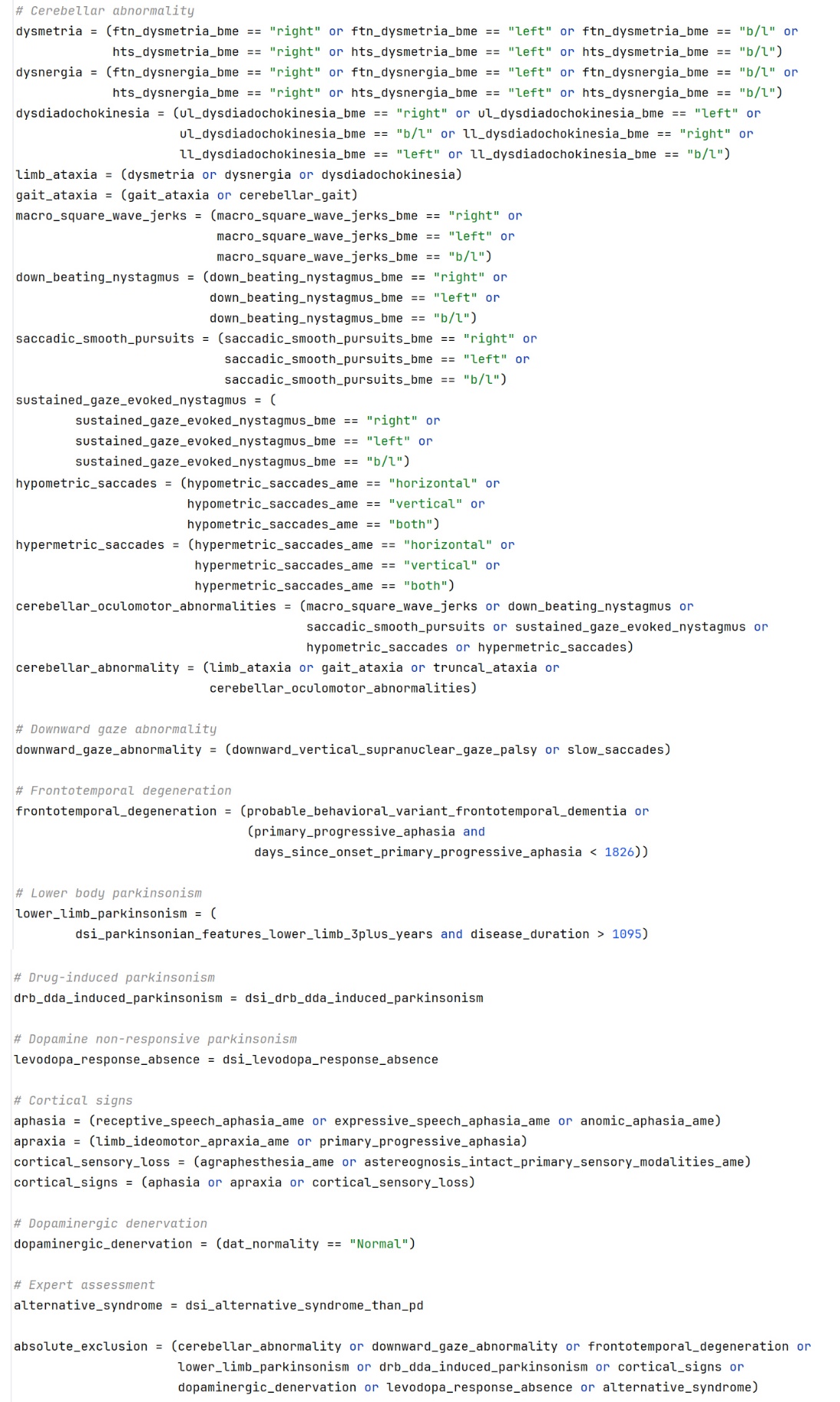
Supplementary Material 4b – Absolute Exclusion Criteria

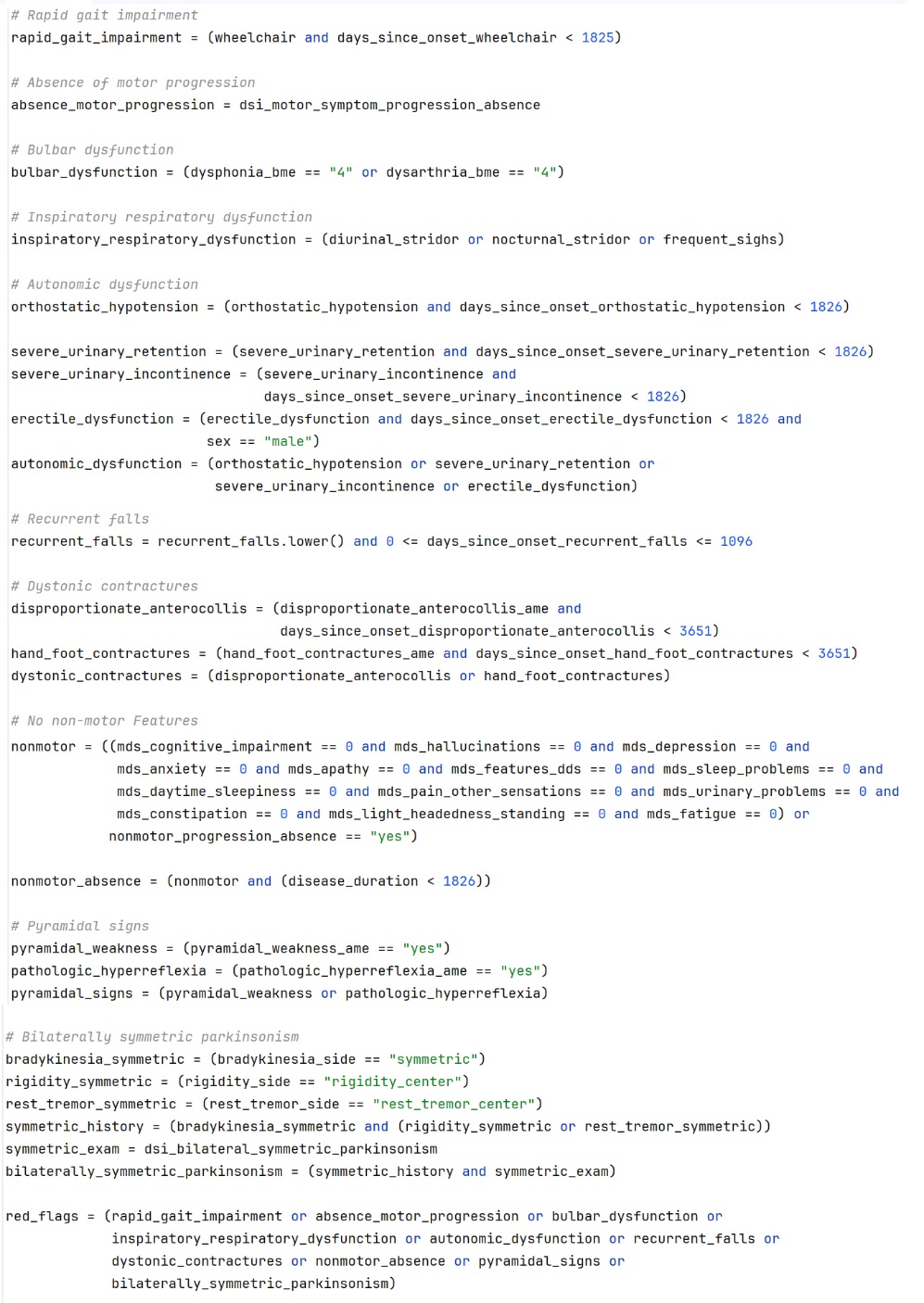
Supplementary Material 4c – Red Flags

Supplementary Material 4d – Supportive Criteria

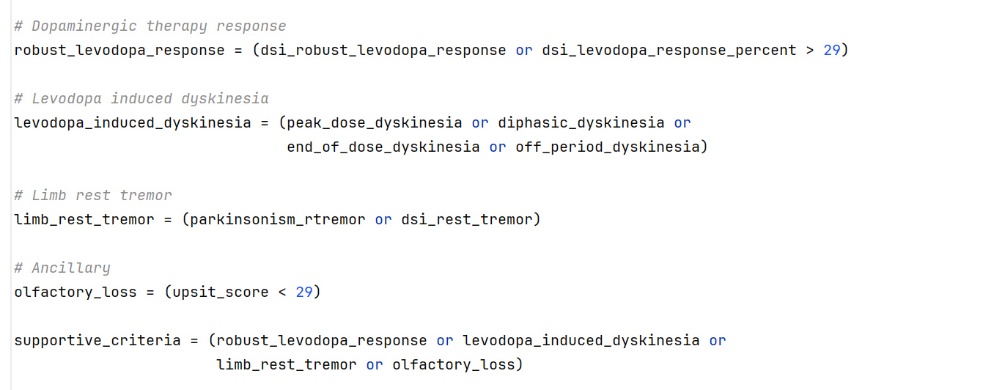
